# Placental SARS-CoV-2 in a patient with mild COVID-19 disease

**DOI:** 10.1101/2020.07.11.20149344

**Authors:** Albert L. Hsu, Minhui Guan, Eric Johannesen, Amanda J. Stephens, Nabila Khaleel, Nikki Kagan, Breanna C. Tuhlei, Xiu-Feng Wan

## Abstract

**Background:** The full impact of COVID-19 on pregnancy remains uncharacterized. Current literature suggests minimal maternal, fetal, and neonatal morbidity and mortality,^1^ and COVID-19 manifestations appear similar between pregnant and non-pregnant women.^2^ We present a case of placental SARS-CoV-2 virus in a woman with an uncomplicated pregnancy and mild COVID-19 disease.

**Methods:** A pregnant woman was evaluated at University of Missouri Women and Children’s Hospital. Institutional review board approval was obtained; information was obtained from medical records. Reverse transcriptase-polymerase chain reaction (RT-PCR) was performed to detect SARS-CoV-2. A gynecological pathologist examined the placenta and performed histolopathology. Sections were formalin-fixed and paraffin-embedded; slides were cut and subjected to hematoxylin-and-eosin or immunohistochemistry (IHC) staining. IHC was performed with specific monoclonal antibodies to detect SARS-CoV-2 antigen or to identify trophoblasts.

**Findings:** A 29 year-old multigravida presented at 40-4/7 weeks for labor induction. With myalgias two days prior, she tested positive for SARS-CoV-2. Her parents were in self-isolation for COVID-19 positivity; husband was asymptomatic and tested negative for COVID-19, but exposed to a workplace (meatpacking facility) outbreak.

Prenatal course was uncomplicated, with no gestational hypertension. She was afebrile and asymptomatic with normal vital signs throughout hospitalization. Her myalgias improved prior to admission. A liveborn male infant was delivered vaginally. Newborn course was uneventful; he was appropriate for gestational age, physical was unremarkable, and he was discharged home at 36 hours. COVID-19 RT-PCR test was negative at 24 hours. At one-week follow-up, newborn was breastfeeding well, with no fevers or respiratory distress.

Overall placental histology is consistent with acute uterine hypoxia (subchorionic laminar necrosis) superimposed on chronic uterine hypoxia (extra-villous trophoblasts and focal chronic villitis). IHC using SARS-CoV-2 nucleocapsid-specific monoclonal antibody demonstrated SARS-CoV-2 antigens throughout the placenta in chorionic villi endothelial cells, and rarely in CK7-expressing trophoblasts. Negative control placenta (November 2019 delivery) and ferret nasal turbinate tissues (not shown) were negative for SARS-CoV-2.

**Interpretation:** In this report, SARS-CoV-2 was found in the placenta, but newborn was COVID-19 negative. Our case shows maternal vascular malperfusion, with no features of fetal vascular malperfusion.

To our knowledge, <u>this is the first report of placental COVID-19 despite</u>*<u>mild</u>*<u>COVID-19 disease</u> in pregnancy (with no symptoms of COVID-19 aside from myalgias); specifically, this patient had no fever, cough, or shortness of breath, but only myalgias and sick contacts. Despite her having mild COVID-19 disease in pregnancy, we demonstrate placental vasculopathy and presence of SARS-CoV-2 virus across the placenta. Evidence of placental COVID-19 raises concern for possible <u>placental vasculopathy</u> (potentially leading to fetal growth restriction, pre-eclampsia, and other pregnancy complications) as well as for <u>potential vertical transmission</u> – especially for pregnant women who may be exposed to COVID-19 in early pregnancy. Further studies are urgently needed, to determine whether women with mild, pre-symptomatic, or asymptomatic COVID-19 may have SARS-CoV-2 virus that can cross the placenta, cause fetal vascular malperfusion, and possibly affect the fetus. This raises important public health and public policy questions of whether future pregnancy guidance should include stricter pandemic precautions, such as screening for a wider array of COVID-19 symptoms, increased antenatal surveillance, and possibly routine COVID-19 testing on a regular basis throughout pregnancy.

## Introduction

As of 6 July 2020, the coronavirus disease 2019 (COVID-19) pandemic has resulted in 2,886,267 cases and 129,811 deaths in the United States.^1^ The full impact of COVID-19 on pregnancy remains to be fully characterized, especially as women who were exposed to COVID-19 early in their first trimester have yet to reach full term, as of the time of writing this manuscript. The current literature suggests minimal maternal, fetal, and neonatal morbidity and mortality for women with COVID-19 in pregnancy.^2^ COVID-19 manifestations have appeared largely similar between pregnant and non-pregnant women,^3^ until a recent publication from the US CDC Morbidity and Mortality Weekly Report (MMWR).^4^

Before this study, we searched PubMed and Medline on 28 May 2020 (updated on 28 June 2020) for articles describing the impact of COVID-19 in pregnancy, using the search terms “COVID-19” or “SARS-CoV-2” and “pregnancy” or “placenta” with no time or language restrictions. We found only previously published research that describes finding SARS-CoV-2 virus in the placentas of women with moderate-to-severe COVID-19 disease. We found no published work about SARS-CoV-2 virus in the placentas of women with mild COVID-19 disease. Two reports suggest that pregnant women with COVID-19 may be more likely to be admitted to the ICU and more likely to be intubated, but all reports consistently state that severe COVID-19 disease during pregnancy is uncommon. We found six manuscripts which describe SARS-CoV-2 virus in the placentas of women with moderate-to-severe COVID-19 disease. We found no published work about SARS-CoV-2 virus immunohistochemistry in the placentas of women with *mild* COVID-19 disease.

In this case study, we present a case of placental SARS-CoV-2 virus in a woman with an uncomplicated pregnancy and mild COVID-19 disease. Here, we review the literature on COVID-19 disease and pregnancy, and subsequently discuss the key messages of this case.

## Case presentation

In April 2020 at the University of Missouri Women and Children’s Hospital, a 29 year-old multigravida presented at 40-4/7 weeks for labor induction. She had tested positive for SARS-CoV-2, with her only symptoms being myalgias two days prior. Her parents and siblings had been in self-isolation for COVID-19 positivity; her husband was asymptomatic and tested negative for COVID-19, but he had been exposed to a workplace (meatpacking facility) outbreak.

Her prenatal course was uncomplicated, with no gestational hypertension or any other pregnancy complications. Her myalgias improved prior to admission. She was afebrile and asymptomatic with normal vital signs throughout hospitalization. Consistent with other reports of laboratory abnormalities associated with COVID-19, her inpatient labwork showed some mild elevations in her LFTs and C-reactive protein, but were otherwise unremarkable (Table 1).

**Table 1.** Significant maternal inpatient laboratory results

| Lab results | Normal values | 4/28/20 | 4/29/20 | 4/30/20 |
| --- | --- | --- | --- | --- |
| C-reactive protein (mg/dL) | 0-0.50 mg/dL | 1.43 mg/dL |  |  |
| D-dimer (mcg/mL) | 0-0.50 mcg/mL | 0.96 mcg/mL | 1.43 mcg/mL | 1.21 mcg/mL |
| Procalcitonin (ng/mL) | 0-0.05 ng/mL | 0.06 ng/mL |  |  |
| Ferritin (ng/mL) | 13.0-150.0 ng/mL |  | 35.9 | 48.3 |
| Alkaline Phosphatase (units/L) | 35-104 units/L | 193 | 175 | 152 |
| AST-SGOT (units/L) | <= 32 units/L | 65 | 57 | 43 |
| ALT-SGPT (units/L) | 10-35 units/L | 214 | 186 | 151 |
| LDH (units/L) | 135-214 units/L | 215 | 226 | 197 |

A liveborn male infant was delivered vaginally. Newborn course was uneventful; with routine resuscitation and newborn APGARs of 8 and 9 at 1 and 5 minutes; points were taken off for color only. Birthweight was 3521g (66^th^ percentile), length was 48 cm (14^th^ percentile) and head circumference was 35 cm (64^th^ percentile), all appropriate for gestational age. His physical was unremarkable, with a small skin tag on his right cheek and dermal melanocytosis on buttock, appropriate for ethnicity. He passed hearing screen and screen for critical congenital heart defects (normal-preductal 99%, postductal 99%.). He was discharged home at 36 hours of life, with a plan for close follow-up from his pediatrician. COVID-19 reverse transcriptase-polymerase chain reaction (RT-PCR) test was negative at 24 hours. At one-week follow-up, newborn was breastfeeding well, with no fevers or respiratory distress. Two months later, mother and baby are doing well.

A gynecological pathologist examined the placenta and performed histolopathology. Sections were formalin-fixed and paraffin-embedded; slides were then cut and subjected to hematoxylin-and-eosin or immunohistochemistry (IHC) staining. Placenta had a marginally-inserted three-vessel umbilical cord; placental membranes were thin and transparent. Placental weight was 538 grams, at the 60^th^ percentile for gestational age. No gross lesions were seen on fetal or maternal placental surfaces; parenchymal sections were unremarkable. On histology, placental membranes showed decidua with scattered arterioles with thickened smooth muscle, consistent with hypertrophic arteriolopathy (Figure 1a) and subchorionic laminar necrosis (1b). Placental disc showed focal lympho-histiocytic inflammation consistent with chronic villitis (1c) and scattered islands of extravillous trophoblasts (1d). No evidence of fetal thrombi (villous sclerosis or villous karyorrhexis) were seen. Overall histology is consistent with acute uterine hypoxia (subchorionic laminar necrosis) superimposed on chronic uterine hypoxia (extra-villous trophoblasts and focal chronic villitis), of which hypertrophic arteriolopathy may be partly responsible.

**Figure 1.**
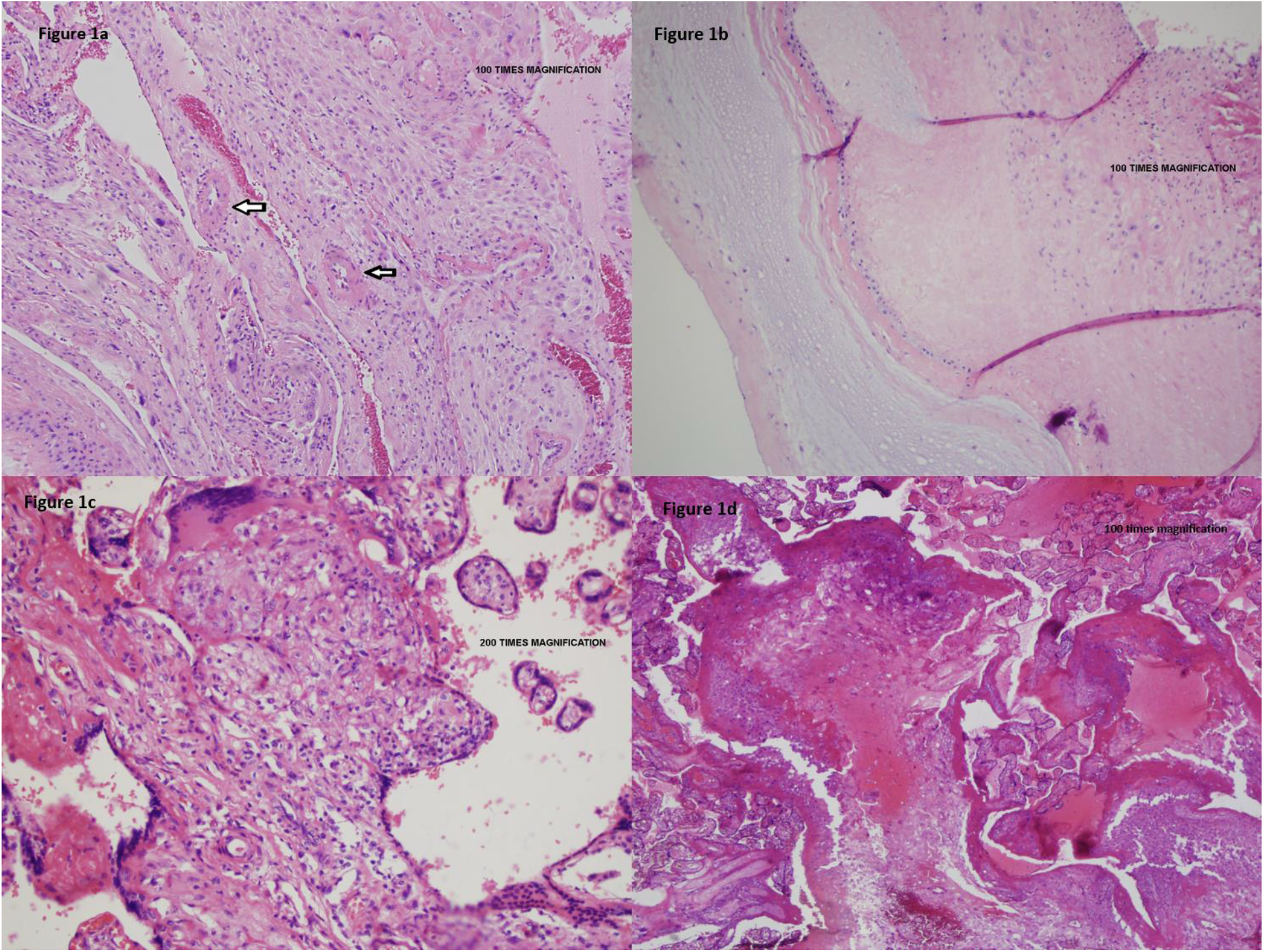
Placental vasculopathy in a pregnant woman with mild COVID-19 disease. Placental membranes showed decidua with scattered arterioles with thickened smooth muscle, consistent with hypertrophic arteriolopathy (vasculopathy) (Figure 1a – umbilical cord and placental membranes) and subchorionic laminar necrosis (Figure 1b – placental parenchyma under the umbilical cord). Placental disc showed focal areas of lympho-histiocytic inflammation consistent with chronic villitis (Figure 1c – central placental parenchyma) and scattered islands of extravillous trophoblasts (Figure 1d – peripheral placental parenchyma).

IHC was performed with SARS-CoV-2 nucelocapsid-specific rabbit monoclonal antibody (Sino Biological, Wayne PA) and goat anti-rabbit IgG (Vector lab, Burlingame, CA). To identify trophoblasts, IHC was performed using rabbit recombinant anti-cytokeratin 7 (CK7) monoclonal antibody (Abcam, Cambridge, MA) and goat anti-rabbit IgG (Vector lab, Burlingame, CA). IHC using SARS-CoV-2 nucleocapsid-specific monoclonal antibody demonstrated SARS-CoV-2 antigens throughout the placenta under the umbilical cord (Figure 2b), and at the central (2c) and peripheral placenta disc (Figure 2d) in chorionic villi endothelial cells, and rarely in CK7-expressing trophoblasts. Negative control placenta (November 2019 delivery, Figure 2a) and ferret nasal turbinate (not shown) were negative for SARS-CoV-2.

**Figure 2.**
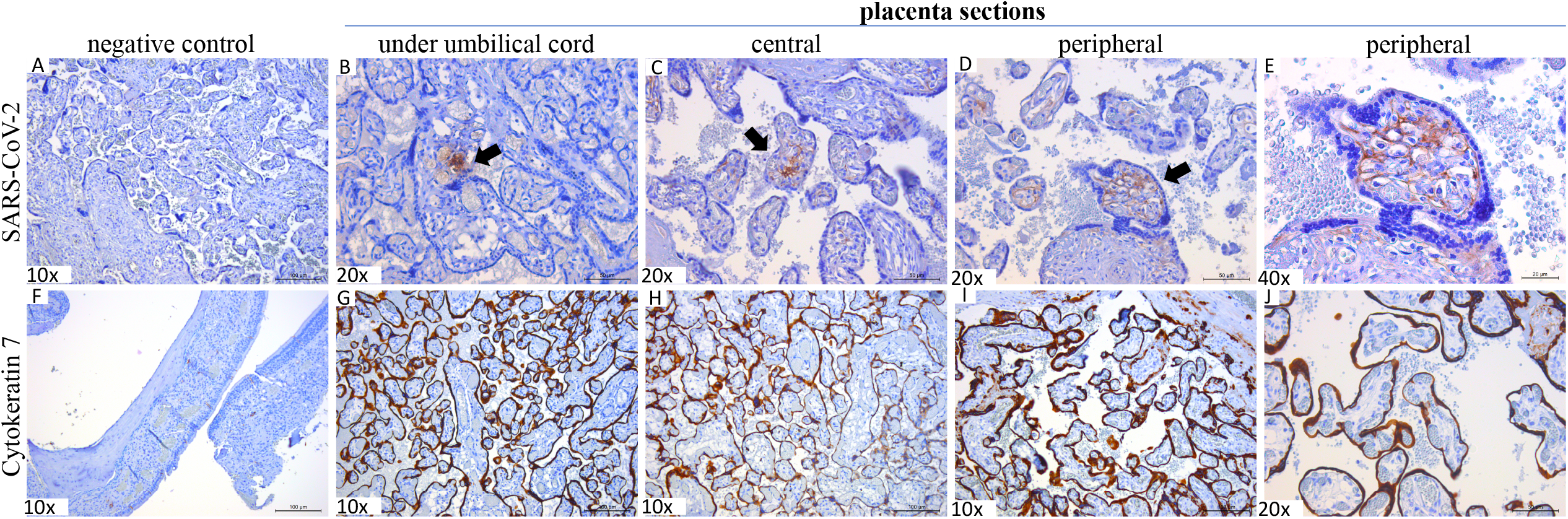
Presence of SARS-CoV-2 virus across the placenta in a patient with mild COVID-19 disease. Immunohistochemistry (IHC) staining of SARS-CoV-2 virus in a COVID-19 negative patient, delivery prior to the COVID-19 outbreak (Figure 2a). IHC of SARS-CoV-2 from three placental sections (2b: under umbilical cord, 2c: central placental disc, 2d-e: peripheral placental disc at 20x and 40x). IHC of cytokeratin-7 (CK-7) marker in control ferret nasal turbinate tissue (Figure 2f). IHC of CK-7 from three placental sections (2g: under umbilical cord, 2h: central placental disc, 2i-j: peripheral placental disc at 10x and 20x). Arrows or brown staining indicate immunoreactive antigens. Bars = 20/50/100 µm shown at the right bottom corner of each panel. IHC was performed with SARS-CoV-2 nucleocapsid-specific rabbit monoclonal antibody (Sino Biological, Wayne, PA) and goat anti-rabbit IgG (Vector lab, Burlingame, CA). To identify trophoblasts, IHC was performed using rabbit recombinant anti-Cytokeratin 7 (CK7) monoclonal antibody (Abcam, Cambridge, MA) and goat anti-rabbit IgG (Vector lab, Burlingame, CA).

## Review

### COVID-19 in pregnancy

Thus far, there have been 12 case series in the literature,^5-16^ which consistently observe that severe COVID-19 disease during pregnancy is uncommon. Out of 431 patients reported in these 12 studies, 36 developed severe or critical disease, and 31 of the 36 severe cases progressed to critical disease and were admitted to the ICU. Overall, however, less than 10% of pregnant patients in these 13 studies are reported to have severe or critical disease.^5-16^ Blitz et al. compared COVID-19 ICU admissions between nonpregnant (n=332, aged 15-49) and pregnant women (n=82) at a large hospital system in New York State^7^ and found that pregnant women were not at an increased risk for ICU admission compared with nonpregnant women (p=0.22);^7^ in this study, 15.1% of nonpregnant females were admitted to the ICU compared to 9.8% of pregnant patients.^7^ Similarly, Savasi et al. reported that out of 77 pregnant women with COVID-19 across 12 hospitals in Italy,^8^ only 6 were admitted to the ICU^8^ and all patients eventually made a complete recovery. Reports of other non-respiratory complications during pregnancy also appear to be rare. A study from Wuhan found that the rate of spontaneous abortion was 12.5% for patients in the first or second trimester (1 out of 8 COVID-19 positive cases), while preeclampsia occurred in 1.5% (1 out of 65 COVID-19 positive cases).^16^ Fortunately, none of these studies reported any maternal deaths.

Two other studies,^4, 17^ suggest that pregnant women with COVID-19 have a higher risk of severe disease than nonpregnant women. A Swedish study showed pregnant Swedish women were 5 times more likely to be admitted to the ICU and 4 times more likely to receive mechanical ventilation than were nonpregnant women.^17^ Also, a US CDC report reviewed 326,335 women of reproductive age (15-44 years of age) with positive SARS-CoV-2 test results.^4^ Data on pregnancy status was available for 28.0% (91,412 women), and 9% of that group (8207 women) were pregnant.^4^ After adjusting for age, race/ethnicity, and underlying conditions, pregnant women were more likely to be admitted to intensive care (aRR = 1.5, 95% confidence interval [CI] = 1.2-1.8) and to receive mechanical ventilation for breathing support (aRR = 1.7, CI 1.2-2.4), but with no significant differences in maternal mortality.^4^ Based on absolute risks, the MMWR report noted that 1.5% of pregnant women with COVID-19 required ICU admission,^4^ so despite the finding of an increased risk of admission to critical care, pregnant women in this study had an even lower overall risk of ICU admission than the previous 12 case series. ^5-16^

All of these studies agree upon a low level of maternal mortality (16 deaths, or 0.2% according to MMWR) from COVID-19.^4-18^ While this is reassuring overall, more data is needed, especially regarding pregnancies with exposure to COVID-19 in the first trimester.

### Possible vertical transmission in pregnancy

There are also several reports suggesting a potential low risk of in utero vertical transmission from mother to fetus. In an Italian series of 22 women affected by COVID-19, Patanè *et al* describe two cases in which the mothers, placentas, and neonates were all positive for SARS-CoV-2 by PCR testing.^19^ Penfield *et al* describe an NYU study of 32 COVID-19 positive pregnant patients, who had 11 placental or membrane swabs sent for analysis, 3 of which were positive for SARS-CoV-2; no infants tested positive for SARS-CoV-2 on days of life 1 through 5, and none demonstrated COVID-19 symptoms.^20^

Baud *et al* describe a second-trimester miscarriage in which the mother (nasopharyngeal swab) and placental submembranes and placental cotyledons were positive for SARS-CoV-2 on RT-PCR.^21^ Hosier *et al* describe SARS-CoV-2 localization to syncytiotrophoblast cells at the maternal-fetal interface of the placenta, with no evidence of vasculopathy.^22^ Chen *et al* describe 9 patients who had a caesarean section, with amniotic fluid, cord blood, neonatal throat swab, and breastmilk samples from six patients all testing negative for SARS-CoV-2.^23^

Ferriaolo *et al* describe an asymptomatic woman with a positive nasopharyngeal swab for COVID-19 in Italy, who had a Caesarean section and positive placental swabs for SARS-CoV-2 RNA.^24^ Dong *et al* report a case of SARS-CoV-2 IgM in a neonate born to a COVID-19 positive mother.^25^ Collectively, this literature suggests that there may be a low risk of vertical transmission of COVID-19 from mother to fetus in utero.

In our report, SARS-CoV-2 was found throughout the placenta on immunohistochemistry, and newborn was COVID-19 negative. As this patient was exposed to COVID-19 in her third trimester of pregnancy, it remains unclear whether women who are exposed to COVID-19 earlier in pregnancy (or multiple times during pregnancy) may have a greater risk of in-utero vertical transmission of COVID-19 from mother to fetus. Further studies are needed, especially from pregnancies with exposure to COVID-19 in the first trimester.

### COVID-19 in the placenta

COVID-19 is known to interact with endothelial cells and cause a hypercoagulative state, which can lead to the formation of microthrombi. ^26^ This can potentially affect the placenta in two ways. First, effects on maternal perfusion of the placenta can manifest as accelerated villous maturation, infarction, intervillous thrombi, extravillous trophoblastic lesions and subchorionic laminar necrosis in the membranes. Secondly, if the fetal circulation is affected, thrombosis of larger vessels, fibrous obliteration of vessels in the villi (villous sclerosis) and breakdown of the endothelial cells within the villous stroma (villous stromal vascular karyorrhexis) may be seen. In a study of placentas from COVID-19 positive mothers in New York and New Jersey, Baergen *et al* showed fetal vascular malperfusion in 8 of 20 placentas and maternal vascular malperfusion in 3 of 20 placentas.^27^ In a study at Northwestern University, Shanes *et al* found that the most common findings in COVID-positive placentas were decidual arteriopathy and intervillous thrombi. ^28^ However, it is notable that signs of fetal and maternal malperfusion are non-specific and can be seen in other conditions, such as hypercoagulable states, like lupus anticoagulant and protein C or S deficiency, gestational hypertension,^29^ pre-eclampsia, as well as in patients with no specific medical history.

## Case Discussion

The inexorable rise in COVID-19 cases in the United States amid decreasing adherence to public health recommendations raises concern for the possibility of viral mutations. In light of the CDC’s MMWR report,^4^ the US CDC also issued updated (6/25/20) recommendations^30^ regarding pregnancy and COVID-19, encouraging pregnant women to “limit interactions with other people as much as possible,” to “take precautions to prevent getting COVID-19 when you do interact with others,” and to be aware that “some babies have tested positive for the virus shortly after birth. It is unknown if these babies got the virus before, during, or after birth.”

While two reports suggest that pregnant women with COVID-19 may be more likely to be admitted to the ICU and more likely to be intubated than non-pregnant women,^4, 17^ but all reports consistently state that severe COVID-19 disease during pregnancy is uncommon.^6-17^ We found six manuscripts which describe SARS-CoV-2 virus in the placentas of women with moderate-to-severe COVID-19 disease. We found no published work about SARS-CoV-2 virus immunohistochemistry in the placentas of women with *mild* COVID-19 disease.

To our knowledge, <u>this is the first report of placental COVID-19 despite</u>*<u>mild</u>*<u>COVID-19 disease</u> in pregnancy (with no symptoms of COVID-19 aside from myalgias); specifically, this patient had no fever, cough, or shortness of breath, but only myalgias and sick contacts. Evidence of placental COVID-19 raises concern for possible <u>placental vasculopathy</u> (potentially leading to fetal growth restriction, pre-eclampsia, and other pregnancy complications) as well as for <u>potential vertical transmission</u> – especially for pregnant women who may be exposed to COVID-19 in early pregnancy. Despite her having mild COVID-19 disease in pregnancy, we demonstrate placental vasculopathy and presence of SARS-CoV-2 virus across the placenta.

While our case shows features of maternal vascular malperfusion (extra-villous trophoblastic lesions and subchorionic laminar necrosis), no evidence of fetal vascular malperfusion were seen. As this patient was exposed to COVID-19 in her third trimester of pregnancy, it remains unclear whether women who are exposed to COVID-19 earlier in pregnancy (or multiple times during pregnancy) may have a greater risk of fetal vascular malperfusion, which may result in pre-eclampsia, fetal growth restriction, or other obstetric complications. Further studies are needed, especially from pregnancies with exposure to COVID-19 in the first trimester.

## Conclusion

To our knowledge, this is the first report of placental COVID-19 despite *mild* COVID-19 disease in pregnancy (with no symptoms of COVID-19 aside from myalgias). All other reports have shown placental COVID-19 in pregnancies with moderate-to-severe disease. This patient had no fever, cough, or shortness of breath, but only myalgias and sick contacts.

In this report, SARS-CoV-2 was found in the placenta, but newborn was COVID-19 negative. Our case shows maternal vascular malperfusion, with no features of fetal vascular malperfusion. Evidence of placental COVID-19 raises concern for placental vasculopathy and potential vertical transmission. Our report raises the question of whether future pregnancy guidance should include even stricter pandemic precautions, such as prenatal screening for a wider array of COVID-19 symptoms, increased antenatal surveillance recommendations, and possibly COVID-19 testing on a regular basis throughout pregnancy.

## Data Availability

Data referred to in the manuscript is shown in tables and figures.

## Author contributions

Drs. Hsu and Wan had full access to all of the data in the study and take responsibility for the integrity of the data and the accuracy of the data analysis.

*Concept and design*: Hsu, Johannesen, Stephens, Khaleel, Wan

Acquisition, analysis, or interpretation of data: Hsu, Guan, Johannesen, Stephens, Khaleel, Tuhlei, Wan

Drafting manuscript: all authors

Critical revision of the manuscript for important intellectual content: all authors

Administrative, technical, or material support: Hsu, Johannesen, Wan

Supervision: Hsu, Wan

## Acknowledgments

Danny Schust, MD; Taylor Nelson, DO; Jane McElroy, PhD; Dima Dandachi, MD; Holly Ford, MD

